# Study on Good Clinical Practices among Researchers in a Tertiary Healthcare Institute in India

**DOI:** 10.1101/2023.07.27.23293264

**Authors:** Harshita, Prasan Kumar Panda

## Abstract

**BACKGROUND:** Good Clinical Practice (GCP) is put in place to protect human participants in clinical trials as well as to ensure the quality of research. Non-adherence to these guidelines can produce research that may not meet the standards set by the scientific community. Therefore, it must be ensured that researchers are well-versed in the GCP. But not much is known about the knowledge and practices of the GCP in the medical colleges of North India.

**AIM:** To assess the knowledge and practices of researchers about GCP and analyze these with respect to the demographics of participants.

**METHODS:** This is a cross-sectional study. A self-structured questionnaire about GCP, after expert validations, was circulated among researchers, at a tertiary healthcare institute, All India Institute of Medical Sciences (AIIMS), Rishikesh. A total of 59 individuals, who were selected by universal sampling, participated in the study. All healthcare workers who have been investigators of Institutional Ethics Committee-approved research projects, except residents and faculty, and are still a part of the institute have been included in the study. The study was approved by the Institutional Ethics Committee of AIIMS, Rishikesh. We used descriptive analysis and the Chi-squared test to analyze data. P-value < 0.05 was considered significant.

**RESULTS:** Out of 59 participants, only 11 (18.6%) were certified for GCP. Most of the participants (64.4%) had “Average” knowledge, 33.9% had “Good” knowledge and 1.7% had “Poor” knowledge. Only 49% of participants had satisfactory practices related to GCP. There was a significant difference in the knowledge based on the current academic position for the items assessing knowledge of institutional review board (P=0.010), confidentiality & privacy (P=0.011), and participant safety & adverse events (P<0.001). There was also a significant difference in knowledge of research misconduct (P=0.024) and participant safety & adverse events (P=0.011) based on certification of GCP. There was a notable difference in the practices related to recruitment & retention on the basis of current academic position (P<0.001) and certification of GCP (P=0.023). We also observed a considerable difference between the knowledge and practices of GCP among the participants (P=0.013).

**CONCLUSION:** Participants have basic knowledge of GCP but show a lack thereof in certain domains of GCP. This can be addressed by holding training sessions focusing on these particular domains.

**Core tip:** There is a lack of knowledge about the Good Clinical Practices in the researchers of medical colleges. In order to improve the quality of research, as well as, make research a better experience for the participants of research, we must work on improving awarenss of the GCP among researchers. This can be done by organising training sessions or workshops which throw light on the principles of GCP.

## INTRODUCTION

With the increase in research studies involving human subjects, there arises a need to have certain guidelines in place to protect human subjects. Additionally, the review of research by the research ethics committee has been mandated by international standards^[1,2]^. A lack of guidelines can lead to misuse of participants as well as other resources. Good Clinical Practice (GCP) provided by the ICH (International Conference on Harmonization) sets ethical and scientific standards and guidelines for conducting research involving human participants^[3]^. The two important principles of these guidelines include protecting the rights of human participants and the credibility of the data generated^[4]^. Such guidelines are required for the welfare of individuals partaking in a trial.

Knowledge of GCP before taking on a research project would lead to increased safety and efficacy of the project. A study conducted in Saudi Arabia, in which 85% of respondents had been trained for GCP, estimated that 97% of respondents believed that GCP guidelines were followed in trials and they improved the quality of the trial. While 59% of the respondents were cynical towards the Institutional Review Board (IRB) approval process or the monitoring of the clinical trials^[5]^. Another study, conducted among dental faculties in India, established that more than 93% of the respondents wanted research ethics education for post-graduates, principal investigators as well as members of the Research Ethics Committee (REC), while less than 20% thought that the REC was not needed because of the presence of scientific committee^[6]^.

The objective of our study was to estimate the knowledge and practices of researchers in a tertiary care institute in Uttarakhand. We also want to determine the difference in knowledge and practices based on research experience, GCP certification, and current academic position in the institute. Our null hypothesis stated that there is no knowledge/practice gap of GCP among researchers in a tertiary care research institution.

## MATERIALS AND METHODS

### Study Design

We conducted a cross-sectional study, which used a self-administered questionnaire at the All India Institute of Medical Sciences, Rishikesh between February and April 2023.

### Study Population

We recruited all the researchers who had Institutional Ethics Committee-approved research projects. Here, the individuals, other than faculty and residents, who have conducted research in the institute are referred to as researchers.

### Questionnaire

The questionnaire was self-structured and based on the GCP guidelines^[7]^. The study tool included 4 sections. The first section contained a summary of the study and informed consent for participation in the study. This was followed by section 2, which had questions that characterized the demographics of the participants. This included age, gender, experience in research, current academic position, number of publications, and certification for GCP. The third section consisted of 13 questions that assessed the knowledge of participants regarding Good Clinical Practice guidelines. 3 of these questions were evaluated by a Likert scale (1-strongly agree, 2- agree, 3- neutral, 4- disagree, 5- strongly disagree), 9 were True/False questions and 1 was a Yes/No question. The last section assessed the application of GCP principles by the participants in practice through 10 case scenarios. In these, 4 options were given out of which one had to be chosen by the participants. Questions were based on the course of Good Clinical Practice by NIDA Clinical Trials Network and references were taken from previous studies^[5,7]^.

All 23 items assessing the Knowledge and practices of participants were categorized into 1 of the 12 modules of NIDA Clinical Trials Network’s course of Good Clinical Practice: introduction (n=2), institutional review boards (n=2), informed consent (n=2), confidentiality & privacy (n=4), participant safety & adverse events (n=3), quality assurance (n=2), the research protocol (n=1), documentation & record-keeping (n=0), research misconduct (n=4), roles & responsibilities (n=0), recruitment & retention (n=2), investigational new drugs (n=1)^[7]^. The questionnaire was subjected to content validation by researchers well versed in the GCP guidelines and modifications were made according to their suggestions. The questionnaire was sent to the participants as Google forms and it was ensured that the participants fully understand the items before responding to them. To ensure quality, the responses were verified and screened for non-response/invalid responses.

### Statistics

Data analysis was done using Jamovi software. We report the positive responses for the items in mean scores.

For True/False and Yes/No items, a score of 1 was given to the correct response and 0 to the incorrect response. For the Likert scale, a score of 0 was given to the most negative response and a score of 4 to the most positive response. The composite score for knowledge was calculated by adding the scores of all 13 items, the maximum possible score was 22. As for the practice-based questions, a score of 1 was given to the correct answer and a score of 0 was given if the participant chose any other options. The total score was calculated by simple summation of the scores of all 10 items (maximum score=10). The knowledge was classified as “good” (score>16), “average” (score= 11-16), and “poor” (score<11). Similarly, the practices of the participants were classified as “satisfactory” (score≥7) and “unsatisfacotry” (score<7).

Demographic data were described using descriptive statistics. Chi-squared test in the bi-variate analysis was used to determine the relationship between knowledge and demographic details like certification of GCP, current academic position, and experience in research. The same was done to determine the association between practices of GCP and independent variables of demographics. To determine the association of knowledge and practices of GCP, a chi-square test was used. P<0.05 was considered significant.

### Ethical Consideration

The approval to conduct this research was taken from the Institutional Ethics Committee of All India Institute of Medical Sciences, Rishikesh, India.

Informed consent, which was subjected to the approval of the Institutional Ethics Committee, was taken from all the individuals before enrolling them in the study.

In order to maintain privacy, any data which may have identified the individual participants of the study was not disclosed.

## RESULTS

A total of 135 individuals were contacted, out of which 67 consented to fill out the questionnaire and 59 responses were valid. Table 1 shows the demographic details of the participants. The age of the respondents ranged from 19 years to 38 years, with a mean age of 23 years and a median of 22 years. Most of the participants were MBBS students(66.1%). Only 10.2% of participants had more than 3 years of research experience while a majority (57.6%) had less than 1 year of experience in conducting a research. 18 individuals (30.5%) had research publications. Out of the respondents, 25.4% have been resource persons for academic classes on research methodologies.

**Table 1:**
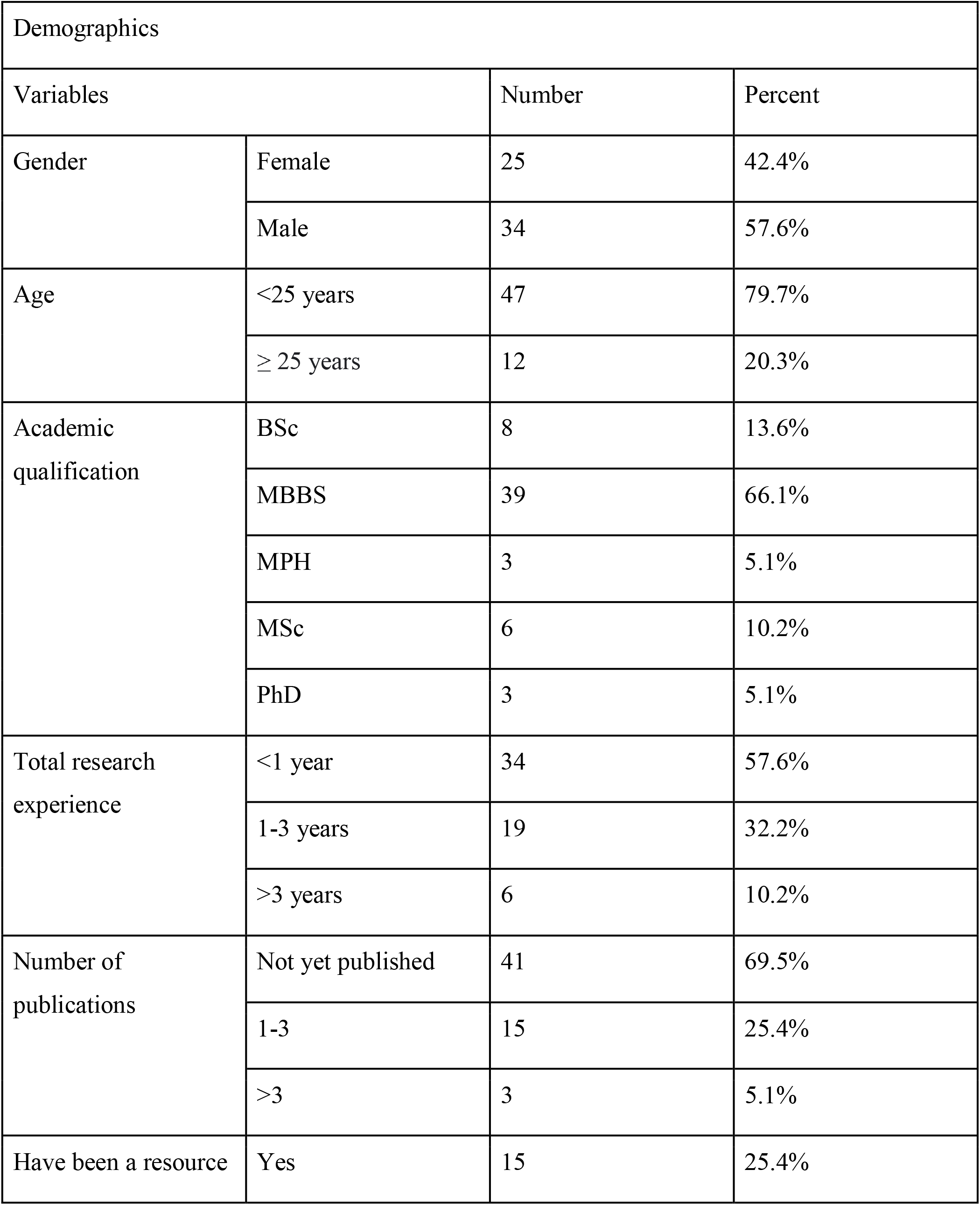
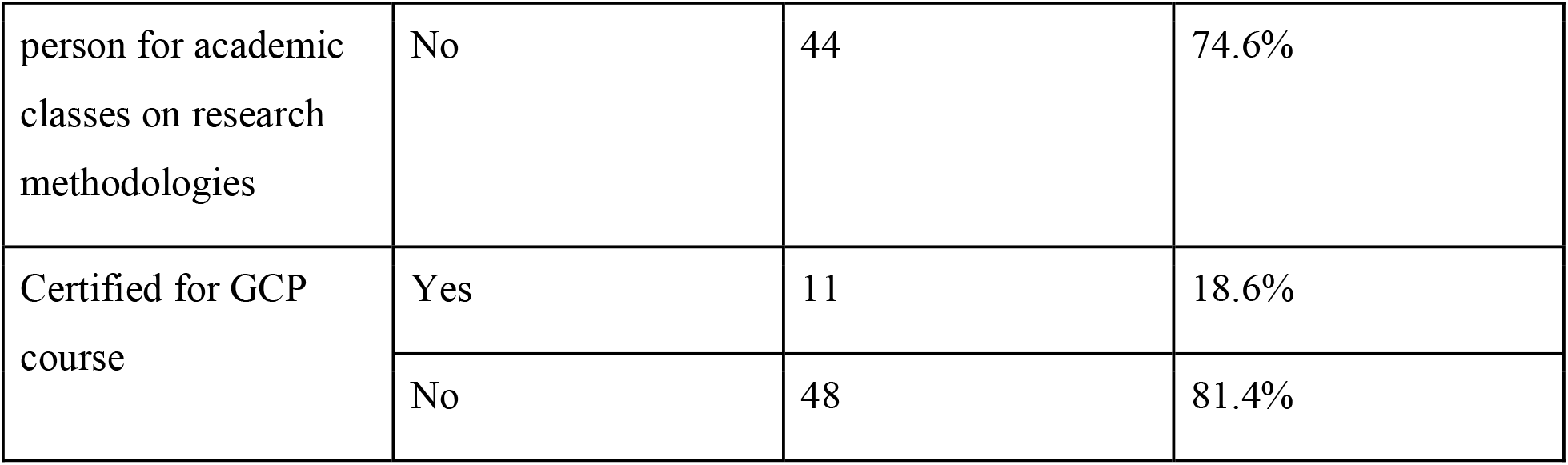
Demographics.

The relationship between research experience and certification of GCP has been shown in Table 2.

**Table 2:** Research Experience with respect to Certification of GCP.

| Certification for GCP | Research experience (in years) | Participants | Percentage of total |
| --- | --- | --- | --- |
| Yes | Less than 1 | 4 | 6.8% |
|  | 1-3 | 4 | 6.8% |
|  | More than 3 | 3 | 5.1% |
| No | Less than 1 | 30 | 50.8% |
|  | 1-3 | 15 | 25.4% |
|  | More than 3 | 3 | 5.1% |

**Knowledge** about GCP

The participants’ knowledge was assessed by 13 items, of which 10 were “Yes/No” or “True/False” items and 3 used the Likert scale. Knowledge scores in our sample ranged from a minimum of 10 (45% correct) to a maximum of 18 (82% correct). The mean score was 15.4 (out of 22) with a standard deviation of 1.95. Knowledge was deemed “Good” if the score was more than equal to 17, “Average” if the score was from 11 to 16, and “Poor” if the score was less than 11.

As seen in Table 4, the maximum number of correct answers were given for the statement “A consent document must be submitted to the Institutional Review Board for its approval before enrolling participants in the study.” (Item no. 2) with 96.6% correct responses. The statement with the maximum number of incorrect answers was “Review by the ethics committee is time consuming and makes it difficult to conduct research.” (Item no. 13) which had 25.4% correct answers.

**Table 4:** Module-wise arrangement of questions assessing Knowledge with % of correct answers.

| Item No. | Percentage of individuals with correct answers |
| --- | --- |
| Introduction |  |
| 1 | 79.7 |
| 12 | 89.8 |
| IRB |  |
| 2 | 96.6 |
| 11 | 94.9 |
| Informed consent |  |
| 3 | 94.9 |
| Confidentiality & Privacy |  |
| 4 | 28.8 |
| 5 | 71.2 |
| Participant safety & adverse events |  |
| 6 | 40.7 |
| 8 | 61.0 |
| Quality assurance |  |
| 7 | 57.6 |
| Research misconduct |  |
| 9 | 57.6 |
| 10 | 37.3 |
| Research protocol |  |
| 13 | 25.4 |

There was a significant difference in the response of the participants on the basis of current academic position for the statements “A consent document must be submitted to the IRB for their approval before enrolling participants in the study.”(p=0.010), “Participant’s information can be disclosed if he/she makes a credible threat to harm another person.” (p=0.011), “The severity of an adverse event is same as its seriousness.” (p<0.001). We also found a considerable difference on the basis of certification of GCP for the following statements: “The severity of an adverse event is same as its seriousness.” (p=0.011), and “Research misconduct consists of-fabrication, falsification and plagiarism. Plagiarism is using another person’s ideas after giving appropriate credit.” (p=0.024)

Most of the respondents (64.4%) had “Average” knowledge and 33.9% had “Good” knowledge. No noteworthy differences in the total knowledge scores were found on the basis of duration of research experience, certification of GCP, and current academic position. This is shown in Table 5.

**Table 5:** Associations between demographics and total knowledge scores.

| Demographics |  | Knowledge of GCP |  |  |  |  |
| --- | --- | --- | --- | --- | --- | --- |
|  |  | Good | Average | Poor | Chi-square | P value |
| Research experience (in years) | < 1 | 13 | 20 | 1 | 1.67 | 0.796 |
|  | 1-3 | 5 | 14 | 0 |  |  |
|  | >3 | 2 | 4 | 0 |  |  |
| Certified for GCP | Yes | 3 | 7 | 1 | 4.54 | 0.103 |
|  | No | 17 | 31 | 0 |  |  |
| Academic position in the institute | BSc | 2 | 6 | 0 | 6.67 | 0.573 |
|  | MBBS | 17 | 21 | 1 |  |  |
|  | MPH | 0 | 3 | 0 |  |  |
|  | MSc | 1 | 5 | 0 |  |  |
|  | PhD | 0 | 3 | 0 |  |  |

### Practices

The practice of the participants in accordance with GCP was assessed with the help of 10 multiple-choice questions, with a score of 1 for each correct response. Out of a total score of 10 for practice-based questions, scores more than equal to 7 were considered “Satisfactory” and scores less than 7 were considered “Unsatisfactory”.

Item numbers 1, 2, and 5 which assess recruitment & retention, informed consent, and research misconduct respectively, had the maximum number of correct responses as shown in Table 6.

**Table 6:** Module-wise arrangement of questions assessing Practices with % of correct answers.

| Item No. | Percentage of individuals with correct answers |
| --- | --- |
| Recruitment & retention |  |
| 1 | 76.3 |
| 3 | 72.9 |
| Informed consent |  |
| 2 | 76.3 |
| Investigational new drugs |  |
| 4 | 52.5 |
| Research misconduct |  |
| 5 | 76.3 |
| 8 | 33.9 |
| Quality assurance |  |
| 6 | 47.5 |
| Confidentiality & privacy |  |
| 7 | 45.8 |
| 10 | 67.8 |
| Participant safety & adverse events |  |
| 9 | 50.8 |

A significant difference was found in the responses for item 3, which assesses the domain of Recruitment & Retention, based on the current academic position (p<0.001) and certification of GCP (p=0.023).

Only 29 participants (49.2%) had “Satisfactory” practices of GCP. Table 7 shows a significant difference in the overall practices of participants on the basis of their current academic position (P=0.008).

**Table 7:** Associations between demographics and total practice scores.

| Demographics |  | Practice of GCP |  |  |  |
| --- | --- | --- | --- | --- | --- |
|  |  | satisfactory | unsatisfactory | Chi-square | P value |
| Research experience (in years) | Less than 1 | 17 | 17 | 0.0357 | 0.982 |
|  | 1-3 | 9 | 10 |  |  |
|  | More than 3 | 3 | 3 |  |  |
| Certified for GCP | Yes | 3 | 8 | 2.59 | 0.108 |
|  | No | 26 | 22 |  |  |
| Academic position in the institute | BSc | 3 | 5 | 13.7 | <sup>a</sup>0.008 |
|  | MBBS | 23 | 16 |  |  |
|  | MPH | 3 | 0 |  |  |
|  | MSc | 0 | 6 |  |  |
|  | PhD | 0 | 3 |  |  |

### Knowledge and Practices

Table 8 shows the results of a chi-squared test for independent samples, which showed a significant relationship between the knowledge and practice scores of the study participants. χ ^2^(2, N=59) =8.62, P= 0.013

**Table 8:** Association between Knowledge and Practices of GCP.

| Practices of GCP | Knowledge of GCP |  |  |
| --- | --- | --- | --- |
|  | Good | Average | Poor |
| Satisfactory | 15 | 14 | 0 |
| Unsatisfactory | 5 | 24 | 1 |
| chi-square | 8.62 |  |  |
| P value | <sup>b</sup>0.013 |  |  |

## DISCUSSION

This questionnaire-based study is the first report in North India specifically targeting knowledge and practices of GCP among researchers (excluding faculty and residents) in a medical college^[5,6,8,9]^.

In our study, most of the researchers were not certified for GCP, which is comparable to a study by Goel D et al. stating a lack of formal training for GCP in health care providers^[8]^. As the research experience increased, more proportion of individuals were certified with GCP. This suggests that individuals with more research experience are more likely to pursue GCP certification. Most of the respondents believed that one must be GCP certified before conducting research, despite this the scores of most of the participants in knowledge and practices were “Average”. This goes on to show that individuals are aware of the GCP on the surface but do not fully understand the principles of GCP. There is a gap in knowledge and practices among participants concerning GCP. The mean score for knowledge of the participants was 70% of the maximum possible score, indicating most of the individuals had some knowledge of GCP. This differs from a study conducted in Japan which showed that ≤ 50% nurses had knowledge about GCP^[10]^. Another study conducted among the doctors in medical colleges of India stated lack of knowledge of GCP^[11]^.

The scores for the knowledge questions related to the topics of IRBs, and informed consent were high, indicating good knowledge of these domains. Participants obtained lower scores for questions related to confidentiality & privacy, research protocol, quality assurance, participant safety and adverse events, and research misconduct, which signifies that participants are less certain or have mixed opinions about these topics. In our study, about 74.6% of the participants believe that the review by the ethics committee is time-consuming or are unsure about this. This is similar to the finding of El-Dessouky et al., Than M M et al.^[9,12]^. This delay could be perceived due to the participants’ lack of understanding of the ethics committee, meticulous evaluation by the ethics committee, or due to the increased workload of the committee.

For the practice-based questions, we observed that statements related to recruitment & retention, and informed consent had a higher mean score in comparison to those related to confidentiality & privacy, quality assurance, participant safety and adverse events, research misconduct, and investigational new drugs.

We also found a significant difference between the knowledge and practices of the study participants for GCP. We also found statistical difference in the knowledge of researchers with respect to certification of GCP, or positions within the institute for items which assessed institutional review board, confidentiality & privacy, participant safety & adverse events, research misconduct. A noteworthy difference was found in the overall practices of GCP based on the academic positions in the institute, along with a difference in practices related to recruitment & retention for various academic positions, and certification of GCP.

At present there is no formal education about Good Clinical Practices in undergraduate or paramedical courses in the institute. In order to build a proper foundation of knowledge of clinical research, one must be familiar with the principles of GCP. We suggest the addition of a course on GCP in the curriculum of undergraduates and post-graduates to familiarize individuals with clinical research. Interactive training sessions can also be held, which have been shown to be effective by some studies^[8,13,14]^.

### Limitations

This study was not able to include the residents as well as faculty of the tertiary care institute as per the directions of the institutional review board. These categories include most of the researchers of the institute, thus their exclusion leading to reduced sample size as well as discrepancies between the study population and the target population. We conducted the research only in a tertiary care institute, which may limit the generalization of our results. Due to fewer participants in the study, there could be a lack in the credibility of subgroup analysis.

## CONCLUSION

Our study concluded that most of the researchers (except faculty and residents) in the medical college are not certified for GCP. Individuals with more research experience are more likely to pursue GCP certification. There was a gap in the knowledge and practices of GCP among the researchers. The understanding of IRB, informed consent, and recruitment & retention was good compared to that of confidentiality & privacy, quality assurance, participant safety and adverse events, and research misconduct. There is a significant difference in knowledge or practices of individuals on the basis of GCP certification or current academic position for some domains of GCP. there is also a significant difference between the knowledge and practices of GCP. This study recommends the incorporation of GCP courses into the academic curriculum or planned training sessions for GCP for the researchers of the institute, which focus on the domains where lack of knowledge was found.

## Data Availability

All data produced in the present study are available upon reasonable request to the authors.

## Institutional review board statement

The study was reviewed and approved by Institutional Ethics Committee of All India Institute of Medical Sciences, Rishikesh, India.

## Informed consent statement

All study participants provided informed written consent before enrolling in the study.

## STROBE statement

The authors have read the STROBE Statement—checklist of items, and the manuscript was prepared and revised according to the STROBE Statement—checklist of items.

